# MODY is prevalent in later-onset diabetes, has potential for targeted therapy but is challenging to identify

**DOI:** 10.1101/2025.06.17.25329143

**Authors:** Luke N Sharp, Uyenlinh L Mirshahi, Kevin Colclough, Timothy S Hall, Jeremy Haley, Stuart Cannon, Thomas W Laver, Michael N Weedon, Andrew T Hattersley, David J Carey, Kashyap A Patel

## Abstract

Maturity Onset Diabetes of the Young (MODY) can present after the age of 40yrs, but its prevalence, clinical characteristics, and the utility of simple clinical features for selecting cases in this age group remain poorly defined. We analysed whole-exome and clinical data from 51,619 individuals with diabetes diagnosed after 40 years of age from two large cohorts: the UK Biobank (n = 25,012) and the US health system MyCode cohort (n = 26,607). The prevalence of MODY due to pathogenic variants in the ten most common genes was 1 in 191 (0.52%) and 1 in 633 (0.16%) in the UK and US cohorts. For subtypes with treatment implications (*GCK, HNF1A, HNF4A, ABCC8, KCNJ11*), prevalence was 1 in 234 and 1 in 935, respectively. *GCK*-MODY was most common, followed by *HNF4A* and lower-penetrance *RFX6*. Clinical features of MODY overlapped with both insulin-treated and non-insulin-treated non-MODY diabetes. Applying simple clinical criteria only increased the MODY diagnosis to 2.64% and 0.87% but missed over 86% of cases. MODY is more common than expected in later-onset diabetes but remains difficult to identify using clinical features alone. Further research is needed to develop more effective strategies for selecting individuals with later-onset diabetes for genetic testing.

**Article Highlights:** *Why did we undertake this study?:* MODY can present later in life, and diagnosis can enable precision treatment. However, individuals with later-onset diabetes are rarely tested.

*What specific question did we ask?:* How common is MODY in people diagnosed with diabetes after 40 years, and can they be identified clinically?

*What did we find?:* MODY affects 1 in 191 to 633 individuals with diabetes onset after 40 years, but clinical features alone cannot reliably identify them.

*What are the implications?:* MODY is relatively common in later-onset diabetes but difficult to detect clinically, limiting routine genetic testing in this group.

## Introduction

Current genetic testing for Maturity Onset Diabetes of the Young (MODY) focuses primarily on individuals diagnosed with diabetes before the age of 40 years (1–4). MODY is a familial form of diabetes caused by heterozygous pathogenic variants in one of 11 genes and typically presents before age 30 (1). This focus on early-onset diabetes reflects the higher prevalence of MODY in younger individuals (∼4%) and the clear clinical benefits of targeted treatment in this group (1; 5). For example, *GCK*-MODY usually requires no pharmacological therapy, while *HNF1A*-, *HNF4A*-, *KCNJ11*-, and *ABCC8*-MODY respond better to sulphonylureas than to insulin (1; 6-8).

It is increasingly recognised that MODY can also present after the age of 40 (9). For example, in a large multigenerational study of *HNF1A*-MODY, approximately 35% of cases developed diabetes after age 40 (10). Identifying these individuals could potentially provide the same benefits from tailored treatment as in younger-onset cases. In younger individuals, clinical features such as age, BMI, parent diabetes and HbA1c help distinguish MODY from more common forms of diabetes and are used in selecting individuals for genetic testing (11). However, it remains unclear whether clinical features can similarly identify MODY in those diagnosed after age 40.

Robust prevalence data and a better understanding of their clinical features are needed before widespread genomic screening for MODY can be considered in this age group. A previous study by Bansal *et al.* investigated genetic aetiologies in this age group but included only 2,670 individuals with diabetes diagnosed after age 40 limiting precision of the estimates (12). Larger studies, such as those by Billings *et al.* (13) and Bonnefond *et al.* (14) , included individuals with diabetes across all ages (n = 14,622 and 25,699, respectively), but did not specifically report on individuals diagnosed after age 40 or provide the clinical features of this group (13; 14). None of these studies assessed whether simple clinical criteria can be used to select individuals for genetic testing in this age group. Therefore, in this study, using whole-exome sequencing and clinical data from 51,619 individuals with diabetes diagnosed after age 40 from two large cohorts, we aimed to determine the prevalence, genetic causes, and clinical features of MODY. We also assessed whether simple clinical criteria could help identify individuals for genetic testing in this age group.

## Research Design and Methods

### Study Population

We used data from the UK Biobank, a population-based cohort of over 500,000 individuals with, whole-exome sequencing, and extensive phenotypic data (15). The dataset includes baseline demographics, age at diabetes diagnosis, and HbA1c measurements, which allow for robust identification of individuals with diabetes (15). For this study, we included 25,012 individuals with diabetes diagnosed at ≥ 40 years of age. The clinical characteristics of these individuals are detailed in Supplementary Table 1. Ethics approval for the UK Biobank study was obtained from the North West Centre for Research Ethics Committee (11/NW/0382). Written informed consent was obtained from all participants.

We also used data from an independent US health system–based Geisinger MyCode cohort of 173,247 individuals from Pennsylvania (16–18). This cohort is enriched for metabolic and cardiovascular diseases and provides an alternatively ascertained cohort more representative of the clinical setting where clinicians may come across these individuals in routine clinical practice (17; 18). This cohort includes detailed genetic data from whole-exome sequencing, along with deep phenotyping derived from electronic health records. We identified 26,607 individuals with diabetes diagnosed ≥ 40 years of age in our study. Supplementary Table 1 provides their clinical characteristics. The US health system–based Geisinger MyCode cohort consisted of individuals who consented to participate in the MyCode Community Health Initiative, an institutional review board approved program to create a biorepository of blood, serum, and DNA samples for broad research use, including genomic analysis. The study was approved as exempt nonhuman research by the Geisinger Institutional Review Board. Deidentified data was analyzed on a subset of the MyCode cohort.

In both cohorts, we defined diabetes based on at least one of the following criteria: self-report by participants, presence of an ICD-9 or ICD-10 code for diabetes, use of diabetes medication, or HbA1c ≥48 mmol/mol prior to recruitment (19). Age at diagnosis was defined as the age at the first recorded evidence of diabetes (19).

### Genetic Data

We used whole-exome sequencing data from the 450,000-participant release of the UK Biobank to identify individuals with MODY. The exome sequencing methodology, including quality control and sample or variant filtering, has been described by Szustakowski *et al.* (20) and is available at: https://biobank.ctsu.ox.ac.uk/showcase/label.cgi?id=170. Individuals from the Geisinger cohort underwent whole-exome sequencing as part of the DiscovEHR collaboration with the Regeneron Genetics Center. The methods for this process have been reported previously by Mirshahi *et al.* (10). We reviewed missense and protein-truncating variants in the ten most common MODY genes, which together account for over 99% of autosomal dominant MODY cases: *ABCC8, GCK, HNF1A, HNF1B, HNF4A, INS, KCNJ11, NEUROD1, PDX1,* and *RFX6* (21). We excluded *CEL* because exome sequencing cannot reliably detect pathogenic variants in this gene (22). We also included *HNF1B* 17q12 deletions in the study as they contribute to >50% of all *HNF1B*-MODY cases (23). We identified individuals with *HNF1B* 17q12 deletions using array data, as described by Cannon *et al.* (24). We included variants classified as pathogenic or likely pathogenic according to ACMG/AMP guidelines and confirmed their quality through manual review of the sequencing read data (25; 26).

## Statistical Analysis

We compared categorical variables using Fisher’s exact test and continuous variables using the Mann–Whitney U test. We calculated exact 95% confidence intervals for proportions and prevalence using the Clopper–Pearson method. We assessed clinical features of individuals with MODY in comparison with two treatment-defined groups: those treated with insulin from diagnosis (defined as starting insulin within one year and remaining on it, with or without adjunctive therapies), and those not treated with insulin at diagnosis. These categories reflect those used in the widely adopted MODY calculator (11; 27). We conducted all analyses using STATA 18 (StataCorp, USA), R, and Python, including the SciPy and statsmodels packages.

## Results

### MODY is prevalent in individuals with diabetes diagnosed after age 40

Among 25,012 UK Biobank participants with diabetes diagnosed after 40 years of age, the prevalence of MODY was 0.52% (1 in 191; 95% CI, 0.44–0.62%; n = 131) (Figure 1A). In the US health system–based Geisinger MyCode cohort of 26,607 individuals with diabetes diagnosed after 40 years of age, the prevalence was lower at 0.16% (1 in 633; 95% CI, 0.11–0.21%; n = 42) (Figure 1A). The combined prevalence of *GCK*, *HNF1A*, *HNF4A*, and *ABCC8*-MODY subtypes with important treatment implications was 0.43% in the UK Biobank (1 in 234; 95% CI, 0.35–0.52%; n = 107) and 0.10% in the US cohort (1 in 985; 95% CI, 0.063–0.14%; n = 27) (Figure 1B). In both cohorts, *GCK* was the most frequent cause, followed by *HNF4A* and the lower-penetrance gene *RFX6* (Figure 1B). All pathogenic variants identified are listed in Supplementary table 2 and 3.

**Figure 1.**
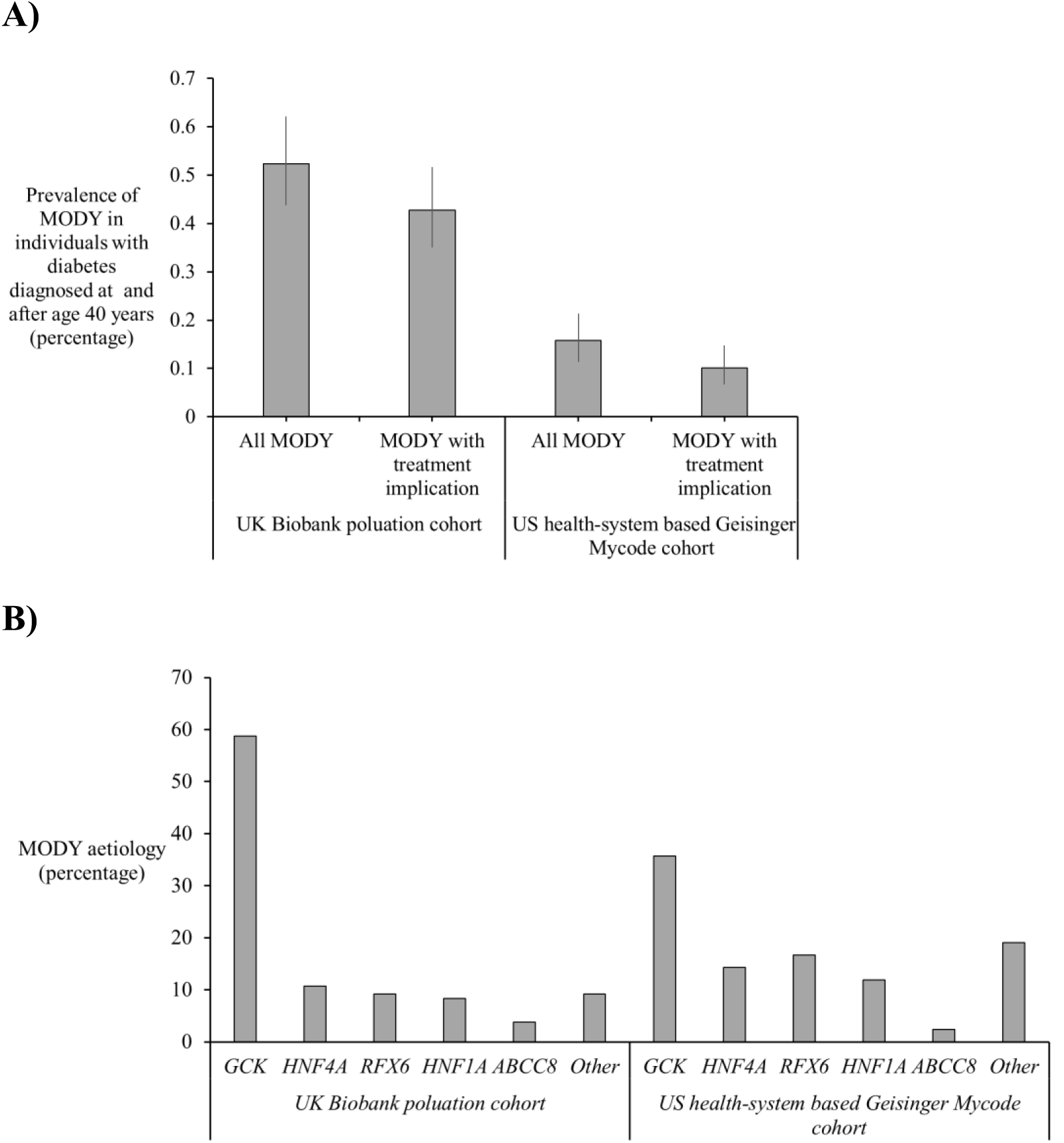
Prevalence and genetic aetiologies of MODY in individuals with diabetes onset at or after 40 years of age. A) Prevalence of MODY among 25,012 diabetes individuals from UK Biobank population cohort and 26,607 individuals from the US health system–based Geisinger MyCode cohort with diabetes diagnosed at after 40 years of age. Prevalence is shown for all MODY genes (pathogenic variants in 10 MODY genes) and for MODY subtypes with treatment-modifying implications (*ABCC8*, *KCNJ11*, *GCK*, *HNF1A*, and *HNF4A*). B) Genetic aetiology of MODY cases in the UK Biobank and MyCode cohort (UK Biobank, *GCK* = 77, *HNF4A* = 14, *RFX6* = 12, *HNF1A* = 11, *ABCC8* = 5, Other (*PDX1*, *NEUROD1*, *HNF1B* = 12. MyCode cohort, *GCK* = 15, *HNF4A* = 6, *RFX6* = 7, *HNF1A* = 5, *ABCC8* = 1, Other (*PDX1*, *HNF1B*= 8)

### Individuals with MODY were on inappropriate treatment for their genotype

Of the 92 individuals with *GCK*-MODY from both cohorts, 37 (40.2%) were receiving pharmacological treatment for diabetes (28 non-insulin and 9 insulin). Of the 42 individuals with *HNF1A*, *HNF4A*, and *ABCC8*-MODY who were on pharmacological diabetes treatment, 73.8% (n = 31) did not receive sulphonylureas (12 non-sulphonylureas, 8 insulin, 11 no pharmacological treatment).

### The clinical features of MODY in individuals diagnosed at and after the age of 40 overlap substantially with those of non-MODY diabetes

To identify features that might guide genetic testing in this age group, we compared individuals with MODY separately against individuals treated with insulin from diagnosis and those not treated with insulin at diagnosis. This approach allowed us to assess which clinical features may support case selection for genetic testing, particularly among insulin-treated individuals, who are likely to benefit most from a genetic diagnosis and targeted therapy. In the UK Biobank, individuals with MODY differed significantly from insulin-treated non-MODY individuals in age at diagnosis, BMI, HbA1c, LDL cholesterol, and parental history of diabetes (Table 1). Among these, HbA1c showed the greatest difference (median 50.6 mmol/mol, IQR 48.5–54.8 vs 59.8, IQR 51.0–69.7; p = 3.6 × 10⁻¹⁵), followed by age at diagnosis (median 54.5 years, IQR 49.4–60.4 vs 50.5, IQR 45.5–57.0; p = 0.00012) (Table 1). We then compared non-insulin-treated at diagnosis individuals to individuals with MODY and found differences in age at diagnosis, BMI, triglycerides, and HDL cholesterol. Of these, BMI and triglycerides showed the most marked differences (median BMI 27.9 kg/m², IQR 25.1–30.2 vs 30.9, IQR 27.8–34.9; p = 6.93×10⁻¹⁴; median triglycerides 1.4 mmol/L, IQR 1.0–2.0 vs 2.0, IQR 1.4–2.8; p = 2.3×10⁻⁹) (Table 1).

**Table 1:** Clinical characteristics of MODY and Non-MODY diabetes in cases with diabetes onset after 40 years in UK Biobank population cohort

| Clinical Features | MODY | Insulin treated from diagnosis | Not insulin treated from diagnosis | P-Value MODY vs insulin treated from diagnosis | P-Value MODY vs not insulin treated from diagnosis |
| --- | --- | --- | --- | --- | --- |
| **N** | 131 | 1,176 | 23,705 | - | - |
| **Age at recruitment, y** | 60.2 (55.1-64.9) | 61.4 (55.8-65.2) | 62.2 (56.8-66.1) | 0.36 | 0.01 |
| **Age at diabetes diagnosis, y** | 54.5 (49.4-60.4) | 50.5 (45.5-57) | 56.5 (50.5-61.5) | 0.00012* | 0.0028* |
| **Females** | 63 (48.1) | 430 (36.6) | 8,896 (37.5) | 0.01 | 0.01 |
| **European Ancestry** | 121 (92.4) | 996 (84.7) | 19,990 (84.3) | 0.02 | 0.01 |
| **Parent with diabetes** | 56 (42.7) | 335 (28.5) | 7,865 (33.2) | 0.0012* | 0.03 |
| **BMI, kg/m<sup>2</sup>** | 27.9 (25.1-30.2) | 29.4 (26.1-33.7) | 30.9 (27.8-34.9) | 0.00014* | 6.93x10<sup>-14</sup>* |
| **HbA1c, mmol/mol** | 50.6 (48.5-54.8) | 59.8 (51-69.7) | 51.1 (46.6-58.3) | 3.6x10<sup>-15</sup>* | 0.76 |
| **Triglycerides, mmol/l** | 1.4 (1-2) | 1.5 (1-2.4) | 2 (1.4-2.8) | 0.45 | 2.3x10<sup>-9</sup>* |
| **HDL, mmol/l** | 1.4 (1-1.6) | 1.2 (1-1.5) | 1.1 (1-1.3) | 0.03 | 1.9x10<sup>-7</sup>* |
| **LDL, mmol/l** | 2.7 (2.3-3.5) | 2.5 (2.1-3) | 2.7 (2.2-3.3) | 0.00039* | 0.7 |
| **Diabetes Treatment** |  |  |  | 3.02x10<sup>-148</sup>* | 0.0032* |
| **Diet/No Treatment** | 73 (55.7) | 0 (0) | 10,752 (45.4) |  |  |
| **Insulin +/- Other** | 16 (12.2) | 1,176 (100) | 1,991 (8.4) |  |  |
| **Non-insulin only** | 42 (32.1) | 0 (0) | 10,962 (46.2) |  |  |

N(%) for categorical data and median (interquartile range) for continuous data. Insulin treated from diagnosis defined as starting insulin within one year and remaining on insulin, with or without adjunctive therapies. \* Below Bonferroni corrected threshold (0.004).

**Table 2:** Clinical characteristics of MODY and Non-MODY diabetes in cases with diabetes onset after 40 years in the US health system–based Geisinger MyCode cohort

| Clinical Features | MODY | Insulin treated from diagnosis | Not insulin treated from diagnosis | P-Value MODY vs insulin treated from diagnosis | P-Value MODY vs not insulin treated from diagnosis |
| --- | --- | --- | --- | --- | --- |
| **N** | 42 | 3,656 | 22,909 | - | - |
| **Age at recruitment, y** | 66.2 (52.8-72.7) | 61.3 (54.3-68.6) | 64.2 (56.8-71.2) | 0.21 | 0.85 |
| **Age at diabetes diagnosis, y** | 57.9 (45.6-65.2) | 56.9 (50.0-64.3) | 58.0 (50.8-65.7) | 0.79 | 0.34 |
| **Females** | 18 (42.9) | 1,846 (50.5) | 11,625 (50.7) | 0.0084 | 0.0089 |
| **European Ancestry** | 42 (100) | 3,302 (90.3) | 21,730 (94.9) | 0.030 | 0.28 |
| **Parent with diabetes** | 22 (52.4) | 1,603 (43.8) | 8,668 (37.8) | 0.53 | 0.057 |
| **BMI, kg/m<sup>2</sup>** | 31.2 (27.4-34.0) | 34.9 (29.8-41.1) | 34.3(29.9-39.8) | 0.0016* | 0.0018* |
| **HbA1c, mmol/mol** | 55.2 (48.6-67.2) | 71.6 (57.4-91.3) | 49.7 (43.2-59.6) | 4.3x10<sup>-6</sup>* | 0.18 |
| **Triglycerides, mmol/l** | 1.7 (0.7-2.1) | 1.9 (1.2-2.9) | 1.9 (1.3-2.7) | 0.22 | 0.25 |
| **HDL, mmol/l** | 1.3 (10.8-1.4) | 1.1 (0.9-1.3) | 1.1 (1-1.3) | 0.00086* | 1.9x10<sup>-7</sup>* |
| **LDL, mmol/l** | 3.0 (2.2-3.5) | 2.6 (1.9-3.3) | 2.7 (2.2-3.4) | 0.23 | 0.63 |
| **Diabetes Treatment** |  |  |  | 8.51x10<sup>-63</sup> | 0.024 |
| **Diet/No Treatment** |  |  |  |  |  |
| **Insulin +/-** | 7 (16.7) | 0 (0) | 5,861 (25.6) |  |  |
| **Others** | 13 (31.0) | 3,656 (100) | 3,488 (15.2) |  |  |
| **Non-insulin only** | 22 (52.4) | 0 (0) | 13,560 (59.2) |  |  |

N(%) for categorical data and median (interquartile range) for continuous data. Insulin treated from diagnosis defined as starting insulin within one year and remaining on insulin, with or without adjunctive therapies. \* Below Bonferroni corrected threshold (0.004).

In the US health system–based Geisinger MyCode which is enriched for metabolic disorders, the clinical features of MODY overlapped even more with those of both non-MODY groups. Only BMI, HbA1c, and HDL cholesterol differed significantly (median BMI 31.2 vs 34.9 vs 34.3 kg/m^2^, median HbA1c 55.2 vs 71.6mmol/mol, median HDL 1.3 vs 1.1 vs 1.1mmol/L), and the magnitude of these differences was small (Table 3). Taken together, these findings suggest that although certain clinical features differ statistically between MODY and non-MODY diabetes after age 40, most differences are modest in clinical terms and show substantial overlap.

**Table 3:**
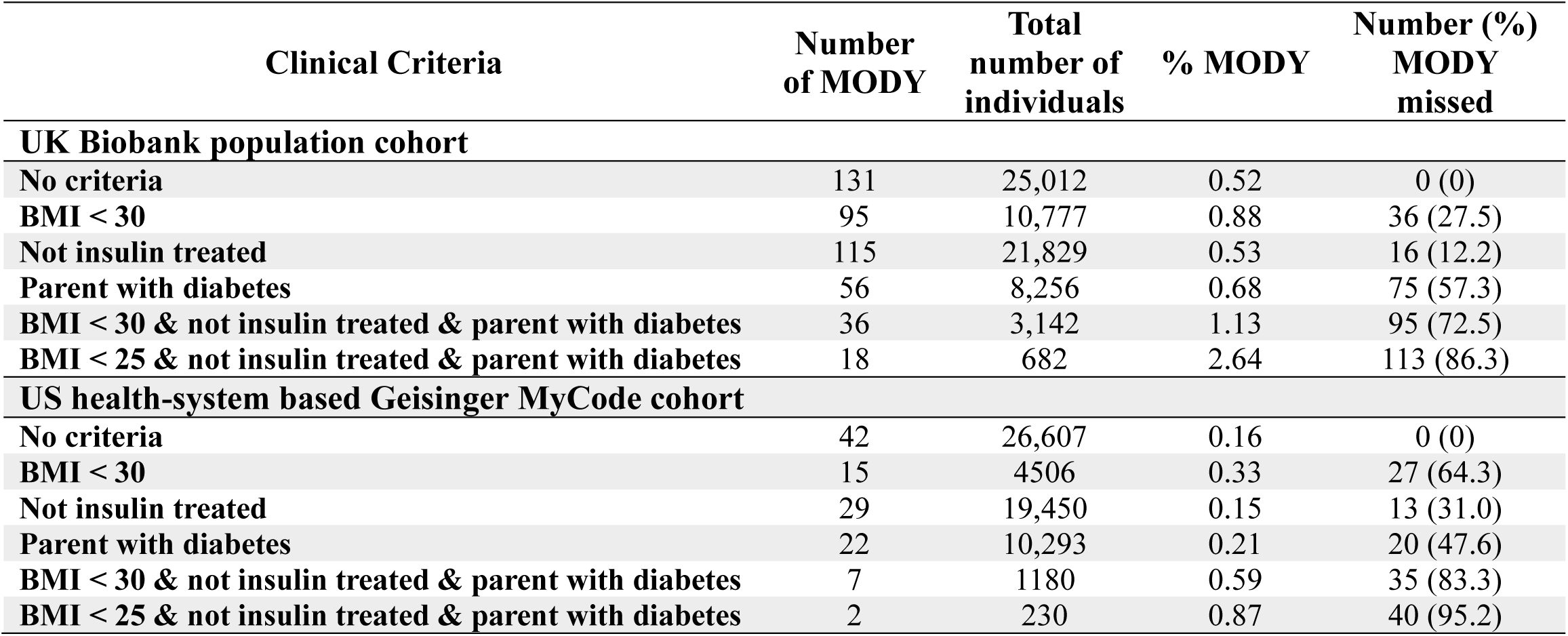
Clinical selection criteria and their utility at identifying individuals with MODY in individuals with diabetes diagnosed after age 40

### Simple clinical criteria alone are insufficient to identify MODY in individuals diagnosed with diabetes at and after the age of 40

We next evaluated a range of clinical criteria, from broad to more stringent, based on classical MODY definitions, to assess their performance in this age group (Table 3). Applying a BMI threshold of <30 kg/m² increased the observed prevalence from 0.52% to 0.88% (95 out of 10,777) in the UK Biobank but missed 27.5% of individuals with MODY. In the US health system–based Geisinger MyCode cohort, the observed prevalence rose from 0.16% to 0.33%, yet 64.3% of individuals with MODY remained undetected. Adding further criteria of no insulin treatment at recruitment and at least one parent with diabetes with BMI raised the observed prevalence to 1.13% (36 out of 3,142) in the UK Biobank and to 0.59% (7 out of 1,180) in the US health system–based Geisinger MyCode cohort. However, these stricter criteria still failed to detect 72.5% and 83.3% of individuals with MODY, respectively.

## Discussion

Our large study of 51,619 individuals with diabetes diagnosed after 40 years of age demonstrates that MODY is relatively common (1 in 191 – 1 in 633) in this age group, has potential for precision therapy but has overlapping clinical features with non-MODY diabetes and it will be difficult to identify the majority of individuals with MODY based on clinical features alone.

MODY prevalence was lower in our US health-system based cohort than in the UK Biobank population cohort. This is likely due to recruitment differences. The UK Biobank has a healthy volunteer bias (15), while the MyCode cohort was enriched for metabolic disease (17; 18), as reflected by a higher average BMI (30.9 vs 33.6kg/m², respectively). These contrasting designs are a strength, enabling prevalence estimates across diverse settings. The true prevalence likely lies between the two estimates (0.16 – 0.52%). Our findings align with Bansal *et al.*, who reported a 0.6% prevalence in this age group (12), and with Gjesing *et al.* (28), who found similar rates of GCK-MODY in late-onset diabetes (UK Biobank:0.31% vs Gjesing *et al.*: 0.8%). Half our cohort was drawn from the UK Biobank, with partial overlap with studies by Billings *et al.* and Bonnefond *et al.* (13; 14). Billings *et al.* analysed 14,622 individuals with diabetes of any age using the 200K exome release from UK Biobank. Bonnefond *et al.* combined 2,151 individuals with diabetes from the UK Biobank (50K release) with 23,548 from other cohorts (n = 25,699). In contrast, we used the 450K exome release and an independent US health system–based cohort, Geisinger Mycode, focusing specifically on diabetes diagnosed after age 40. This targeted approach offers a largely non-overlapping, age-specific view of MODY in later-onset diabetes.

MODY is less common in individuals diagnosed after age 40 than in those with young-onset diabetes and has distinctly different genetic causes. The prevalence of MODY in early-onset diabetes is approximately 4%, significantly higher than individuals with later-onset diabetes (5). This difference reflects the distinct genetic aetiologies in these groups. For example, variants in the highly penetrant *HNF1A* gene are the most common cause of progressive diabetes, but it ranked fourth among those diagnosed after age 40 in the UK Biobank and MyCode cohort (21; 29). In contrast to individuals with early-onset MODY, late-onset MODY includes a higher proportion of lower-penetrance genes. *RFX6*, *NEUROD1*, and *PDX1*-MODY together accounted for 15.3% of late-onset MODY in the UK Biobank, compared to only 4.2% in young-onset cases (21). *GCK*-MODY is an exception, with similar prevalence across age groups. This reflects its distinct pathophysiology: *GCK*-MODY causes mild, lifelong fasting hyperglycaemia that begins at birth and gradually worsens with age (7; 30; 31). As a result, age at diagnosis reflects when hyperglycaemia is detected rather than when the variant becomes penetrant. Alongside increased diabetes screening in older adults, this explains the consistent frequency of *GCK*-MODY across age groups.

Our results show substantial overlap in clinical features between MODY and other forms of diabetes diagnosed after age 40, making clinical recognition difficult. Combined with its lower prevalence in this age group, this limits the ability to select individuals with sufficient prior probability for routine genetic testing. For example, in young-onset referrals, clinical suspicion yields a MODY positivity rate of approximately 20% (21). In contrast, even under ideal criteria in individuals with diabetes diagnosed after 40 years of age (BMI < 25, not insulin-treated, and with a parent affected by diabetes), the positivity rate was only 1–2.6%, and 87% of the individuals with MODY would be missed. Adding biomarkers such as islet autoantibodies or C-peptide in insulin-treated patients may improve detection (1). However, the overall low yield, predominance of low-penetrance variants, and limited impact on treatment decisions argue against routine genomic screening for MODY in this age group. Instead, genetic testing can be considered on a case-by-case basis, where it is supported by additional evidence—for example, a younger affected family member or specific clinical features suggesting an underlying aetiology, such as renal cysts in *HNF1B*-MODY (22).

Our study has several limitations. We lacked data on type 1 diabetes biomarkers, such as islet autoantibodies and C-peptide. Both factors would have help selecting individuals for MODY testing. Although we included participants of all ethnicities, the cohorts were predominantly of European ancestry, which may limit generalisability. We focused on the ten most common MODY genes and did not assess *CEL* or mitochondrial diabetes caused by the m.3243A>G variant, so our prevalence estimates may be slightly conservative. Rare syndromic or recessive forms of monogenic diabetes were also excluded, though these account for less than 7% of monogenic diabetes cases and are unlikely to substantially alter overall prevalence estimates (21). Finally, we restricted our analysis to genes with definite or limited evidence of causality according to ClinGen (32) and did not include refuted genes.

In summary, MODY is present in individuals diagnosed with diabetes after age 40, but its genetic aetiology differs from that seen in younger-onset cases. The substantial overlap in clinical features with other forms of later-onset diabetes makes implementing routine genomic screening in this group challenging.

## Article Information

## Acknowledgements

The authors are grateful for the use of the genomic and clinical data from the MyCode participants. The patient enrolment and exome sequencing were funded by the Regeneron Genetics Center. We are grateful to the Geisinger-Regeneron DiscovEHR collaboration for making the genotype and phenotype data available.

This research was conducted using the UK Biobank Resource. This work was conducted under UK Biobank project number 103356. The current work is funded by Diabetes UK (19/0005994 and 21/0006335), MRC (MR/T00200X/1). KAP is funded by the Wellcome Trust (219606/Z/19/Z).

T.W.L is supported by the Academy of Medical Sciences/the Wellcome Trust/the Government Department of Science Innovation and Technology/the British Heart Foundation/Diabetes UK Springboard Award [SBF009\1135]. The work is supported by the National Institute for Health Research (NIHR) Exeter Biomedical Research Centre, and Clinical Research facilities, Exeter, UK. The Wellcome Trust, MRC and NIHR had no role in the design and conduct of the study; collection, management, analysis, and interpretation of the data; preparation, review, or approval of the manuscript; and decision to submit the manuscript for publication. The views expressed are those of the author(s) and not necessarily those of the Wellcome Trust, Department of Health, NHS or NIHR.

For the purpose of open access, the author has applied a CC BY public copyright licence to any Author Accepted Manuscript version arising from this submission.

## Data availability

The UK Biobank data used in this study is freely accessible from the UK Biobank https://www.ukbiobank.ac.uk. Data from the Geisinger MyCode cohort are available within the article and its supplemental information. Additional information is available upon request, subject to data user agreement.

## Author Contributions

L.N.S and U.L.M researched data, contributed to discussion and wrote the first draft of the manuscript. K.C, J.H and S.C ,T.S.H, T.W.L, M.N.W, A.T.H and D.J.C, researched data, contributed to discussion, and edited the paper. K.A.P is the guarantor of this work.

## References

1. Colclough K, Patel K: How do I diagnose Maturity Onset Diabetes of the Young in my patients? Clin Endocrinol (Oxf) 2022;97:436–447

2. Harsunen M, Kettunen JLT, Härkönen T, Dwivedi O, Lehtovirta M, Vähäsalo P, Veijola R, Ilonen J, Miettinen PJ, Knip M, Tuomi T: Identification of monogenic variants in more than ten per cent of children without type 1 diabetes-related autoantibodies at diagnosis in the Finnish Pediatric Diabetes Register. Diabetologia 2023;66:438–449

3. Irgens HU, Molnes J, Johansson BB, Ringdal M, Skrivarhaug T, Undlien DE, Søvik O, Joner G, Molven A, Njølstad PR: Prevalence of monogenic diabetes in the population-based Norwegian Childhood Diabetes Registry. Diabetologia 2013;56:1512–1519

4. Johnson SR, Ellis JJ, Leo PJ, Anderson LK, Ganti U, Harris JE, Curran JA, McInerney-Leo AM, Paramalingam N, Song X, Conwell LS, Harris M, Jones TW, Brown MA, Davis EA, Duncan EL: Comprehensive genetic screening: The prevalence of maturity-onset diabetes of the young gene variants in a population-based childhood diabetes cohort. Pediatr Diabetes 2019;20:57–64

5. Shields BM, Shepherd M, Hudson M, McDonald TJ, Colclough K, Peters J, Knight B, Hyde C, Ellard S, Pearson ER, Hattersley AT: Population-Based Assessment of a Biomarker-Based Screening Pathway to Aid Diagnosis of Monogenic Diabetes in Young-Onset Patients. Diabetes Care 2017;40:1017–1025

6. Hattersley AT, Patel KA: Precision diabetes: learning from monogenic diabetes. Diabetologia 2017;60:769–777

7. Steele AM, Shields BM, Wensley KJ, Colclough K, Ellard S, Hattersley AT: Prevalence of vascular complications among patients with glucokinase mutations and prolonged, mild hyperglycemia. Jama 2014;311:279–286

8. Thuesen ACB, Jensen RT, Maagensen H, Kristiansen MR, Sørensen HT, Vaag A, Beck-Nielsen H, Pedersen OB, Grarup N, Nielsen JS, Rungby J, Gjesing AP, Storgaard H, Vilsbøll T, Hansen T: Identification of pathogenic GCK variants in patients with common type 2 diabetes can lead to discontinuation of pharmacological treatment. Mol Genet Metab Rep 2023;35:100972

9. Owen KR: Monogenic diabetes in adults: what are the new developments? Curr Opin Genet Dev 2018;50:103–110

10. Kettunen JLT, Rantala E, Dwivedi OP, Isomaa B, Sarelin L, Kokko P, Hakaste L, Miettinen PJ, Groop LC, Tuomi T: A multigenerational study on phenotypic consequences of the most common causal variant of HNF1A-MODY. Diabetologia 2022;65:632–643

11. Shields BM, Carlsson A, Patel K, Knupp J, Kaur A, Johnston D, Colclough K, Larsson HE, Forsander G, Samuelsson U, Hattersley A, Ludvigsson J: Development of a clinical calculator to aid the identification of MODY in pediatric patients at the time of diabetes diagnosis. Sci Rep 2024;14:10589

12. Bansal V, Gassenhuber J, Phillips T, Oliveira G, Harbaugh R, Villarasa N, Topol EJ, Seufferlein T, Boehm BO: Spectrum of mutations in monogenic diabetes genes identified from high-throughput DNA sequencing of 6888 individuals. BMC Med 2017;15:213

13. Billings LK, Shi Z, Resurreccion WK, Wang CH, Wei J, Pollin TI, Udler MS, Xu J: Statistical evidence for high-penetrance MODY-causing genes in a large population-based cohort. Endocrinol Diabetes Metab 2022;5:e372

14. Bonnefond A, Boissel M, Bolze A, Durand E, Toussaint B, Vaillant E, Gaget S, Graeve F, Dechaume A, Allegaert F, Guilcher DL, Yengo L, Dhennin V, Borys JM, Lu JT, Cirulli ET, Elhanan G, Roussel R, Balkau B, Marre M, Franc S, Charpentier G, Vaxillaire M, Canouil M, Washington NL, Grzymski JJ, Froguel P: Pathogenic variants in actionable MODY genes are associated with type 2 diabetes. Nat Metab 2020;2:1126–1134

15. Bycroft C, Freeman C, Petkova D, Band G, Elliott LT, Sharp K, Motyer A, Vukcevic D, Delaneau O, O’Connell J, Cortes A, Welsh S, Young A, Effingham M, McVean G, Leslie S, Allen N, Donnelly P, Marchini J: The UK Biobank resource with deep phenotyping and genomic data. Nature 2018;562:203–209

16. Savatt JM, Kelly MA, Sturm AC, McCormick CZ, Williams MS, Nixon MP, Rolston DD, Strande NT, Wain KE, Willard HF, Faucett WA, Ledbetter DH, Buchanan AH, Martin CL: Genomic Screening at a Single Health System. JAMA Netw Open 2025;8:e250917

17. Carey DJ, Fetterolf SN, Davis FD, Faucett WA, Kirchner HL, Mirshahi U, Murray MF, Smelser DT, Gerhard GS, Ledbetter DH: The Geisinger MyCode community health initiative: an electronic health record-linked biobank for precision medicine research. Genet Med 2016;18:906–913

18. Dewey FE, Murray MF, Overton JD, Habegger L, Leader JB, Fetterolf SN, O’Dushlaine C, Van Hout CV, Staples J, Gonzaga-Jauregui C, Metpally R, Pendergrass SA, Giovanni MA, Kirchner HL, Balasubramanian S, Abul-Husn NS, Hartzel DN, Lavage DR, Kost KA, Packer JS, Lopez AE, Penn J, Mukherjee S, Gosalia N, Kanagaraj M, Li AH, Mitnaul LJ, Adams LJ, Person TN, Praveen K, Marcketta A, Lebo MS, Austin-Tse CA, Mason-Suares HM, Bruse S, Mellis S, Phillips R, Stahl N, Murphy A, Economides A, Skelding KA, Still CD, Elmore JR, Borecki IB, Yancopoulos GD, Davis FD, Faucett WA, Gottesman O, Ritchie MD, Shuldiner AR, Reid JG, Ledbetter DH, Baras A, Carey DJ: Distribution and clinical impact of functional variants in 50,726 whole-exome sequences from the DiscovEHR study. Science 2016;354

19. Mirshahi UL, Colclough K, Wright CF, Wood AR, Beaumont RN, Tyrrell J, Laver TW, Stahl R, Golden A, Goehringer JM, Frayling TF, Hattersley AT, Carey DJ, Weedon MN, Patel KA: Reduced penetrance of MODY-associated HNF1A/HNF4A variants but not GCK variants in clinically unselected cohorts. Am J Hum Genet 2022;109:2018–2028

20. Szustakowski JD, Balasubramanian S, Kvikstad E, Khalid S, Bronson PG, Sasson A, Wong E, Liu D, Wade Davis J, Haefliger C, Katrina Loomis A, Mikkilineni R, Noh HJ, Wadhawan S, Bai X, Hawes A, Krasheninina O, Ulloa R, Lopez AE, Smith EN, Waring JF, Whelan CD, Tsai EA, Overton JD, Salerno WJ, Jacob H, Szalma S, Runz H, Hinkle G, Nioi P, Petrovski S, Miller MR, Baras A, Mitnaul LJ, Reid JG: Advancing human genetics research and drug discovery through exome sequencing of the UK Biobank. Nat Genet 2021;53:942–948

21. Colclough K, Ellard S, Hattersley A, Patel K: Syndromic Monogenic Diabetes Genes Should Be Tested in Patients With a Clinical Suspicion of Maturity-Onset Diabetes of the Young. Diabetes 2022;71:530–537

22. El Jellas K, Dušátková P, Haldorsen IS, Molnes J, Tjora E, Johansson BB, Fjeld K, Johansson S, Průhová Š, Groop L, Löhr JM, Njølstad PR, Molven A: Two New Mutations in the CEL Gene Causing Diabetes and Hereditary Pancreatitis: How to Correctly Identify MODY8 Cases. J Clin Endocrinol Metab 2022;107:e1455–e1466

23. Sánchez-Cazorla E, Carrera N, García-González M: HNF1B Transcription Factor: Key Regulator in Renal Physiology and Pathogenesis. Int J Mol Sci 2024;25

24. Cannon S, Clissold R, Sukcharoen K, Tuke M, Hawkes G, Beaumont RN, Wood AR, Gilchrist M, Hattersley AT, Oram RA, Patel K, Wright C, Weedon MN: Recurrent 17q12 microduplications contribute to renal disease but not diabetes. J Med Genet 2023;60:491–497

25. ACGS Best Practice Guidelines for Variant Classification in Rare Disease 2020 [article online], 2020. Available from https://www.acgs.uk.com/media/11631/uk-practice-guidelines-for-variant-classification-v4-01-2020.pdf.

26. Robinson JT, Thorvaldsdóttir H, Winckler W, Guttman M, Lander ES, Getz G, Mesirov JP: Integrative genomics viewer. Nat Biotechnol 2011;29:24–26

27. Shields BM, McDonald TJ, Ellard S, Campbell MJ, Hyde C, Hattersley AT: The development and validation of a clinical prediction model to determine the probability of MODY in patients with young-onset diabetes. Diabetologia 2012;55:1265–1272

28. Gjesing AP, Engelbrechtsen L, Cathrine BTA, Have CT, Hollensted M, Grarup N, Linneberg A, Steen Nielsen J, Christensen LB, Thomsen RW, Johansson KE, Cagiada M, Gersing S, Hartmann-Petersen R, Lindorff-Larsen K, Vaag A, Sørensen HT, Brandslund I, Beck-Nielsen H, Pedersen O, Rungby J, Hansen T: 14-fold increased prevalence of rare glucokinase gene variant carriers in unselected Danish patients with newly diagnosed type 2 diabetes. Diabetes Res Clin Pract 2022;194:110159

29. Elashi AA, Toor SM, Diboun I, Al-Sarraj Y, Taheri S, Suhre K, Abou-Samra AB, Albagha OME: The Genetic Spectrum of Maturity-Onset Diabetes of the Young (MODY) in Qatar, a Population-Based Study. Int J Mol Sci 2022;24

30. Steele AM, Wensley KJ, Ellard S, Murphy R, Shepherd M, Colclough K, Hattersley AT, Shields BM: Use of HbA1c in the identification of patients with hyperglycaemia caused by a glucokinase mutation: observational case control studies. PLoS One 2013;8:e65326

31. Kentistou KA, Lim BEM, Kaisinger LR, Steinthorsdottir V, Sharp LN, Patel KA, Tragante V, Hawkes G, Gardner EJ, Olafsdottir T, Wood AR, Zhao Y, Thorleifsson G, Day FR, Ozanne SE, Hattersley AT, O’Rahilly S, Stefansson K, Ong KK, Beaumont RN, Perry JRB, Freathy RM: Rare variant associations with birth weight identify genes involved in adipose tissue regulation, placental function and insulin-like growth factor signalling. Nat Commun 2025;16:648

32. Rehm HL, Berg JS, Brooks LD, Bustamante CD, Evans JP, Landrum MJ, Ledbetter DH, Maglott DR, Martin CL, Nussbaum RL, Plon SE, Ramos EM, Sherry ST, Watson MS: ClinGen--the Clinical Genome Resource. N Engl J Med 2015;372:2235–2242

