## Supplemental Tables 1, 2 and 3 for "MODY is prevalent in later-onset diabetes, has potential for targeted therapy but is challenging to identify"

### Supplementary Materials

**Supplementary Table 1: Clinical characteristics of individuals diagnosed with diabetes after age 40yrs in the UK Biobank and MyCode population.**

| Clinical Features | UK Biobank | MyCode |
|---|---|---|
| **N** | 25,012 | 26,607 |
| **Age at recruitment, y** | 62.2 (56.8-66.1) | 63.9 (56.2-71.3) |
| **Age at diagnosis, y** | 56.5 (50.5-61.5) | 57.9 (50.7-65.5) |
| **Females** | 9,389 (37.5%) | 13,489 (50.7) |
| **European Ancestry** | 21,107 (84.4%) | 25,073 (94.2) |
| **Parent with diabetes** | 8,256 (33.0%) | 10,293 (38.7) |
| **BMI, Kg/m$^2$** | 30.9 (27.7-34.8) | 33.6 (29.1-38.6) |

N (%) for categorical data and median (interquartile range) for the continuous data.

**Supplemental Table 2: Pathogenic variants from the UK Biobank identified in this study.**

| Genomic Change | Gene | Protein Change | Nucleotide Change | Criteria | Classification |
|---|---|---|---|---|---|
| 11:17397788:C:T | ABCC8 | p.Gly1256Ser | ENST00000302539.9:c.3766G>A | PM2 PP3 PM6 PS4_Moderate PM3 PP4_Moderate | Likely Pathogenic |
| 11:17460529:C:T | ABCC8 | p.Val324Met | ENST00000302539.9:c.970G>A | PM2 PP3 PS4_Strong PP4_Moderate PM3 PS2 | Pathogenic |
| 11:17404525:G:A | ABCC8 | p.Arg1183Trp | ENST00000302539.9:c.3547C>T | PM2 PP3 PS4_Strong PP4_Moderate PM3 PM5 PP1_Strong PS2 | Pathogenic |
| 11:17413396:G:A | ABCC8 | p.Arg826Trp | ENST00000302539.9:c.2476C>T | PM2 PP3 PS4_Strong PP4_Moderate PM3 PP1_Strong | Pathogenic |
| 11:17460582:C:T | ABCC8 | p.Arg306His | ENST00000302539.9:c.917G>A | PM2 PP3 PS4 PP4_Moderate PM3 | Pathogenic |
| 7:44149977:G:A | GCK | p.Arg191Trp | ENST00000403799.8:c.571C>T | PS4 PS3 PP1_Strong PM6 PP4_Moderate PM2 PP2 PP3 | Pathogenic |
| 7:44151069:C:T | GCK | p.Asp124Asn | ENST00000403799.8:c.370G>A | PM2 PP3 PP1_Strong PS4_Moderate PM5_Supporting PP4 PP2 | Pathogenic |
| 7:44150004:C:T | GCK | p.Val182Met | ENST00000403799.8:c.544G>A | PM2 PP1_Strong PS4 PM5_Supporting PP4 PP2 PS3 | Pathogenic |
| 7:44149797:G:GT | GCK | p.Tyr214* | ENST00000403799.8:c.641dup | PVS1 PM2 PS4_Supporting PP1_Strong PP4 | Pathogenic |
| 7:44149860:C:T | GCK | p.? | ENST00000403799.8:c.580-1G>A | PVS1 PM2 PP3 PS4_Supporting PP4 | Pathogenic |
| 7:44153326:G:T | GCK | p.Tyr61* | ENST00000403799.8:c.183C>A | PVS1 PM2 PP1_Strong PS4 PP4 | Pathogenic |
| 7:44145522:C:G | GCK | p.Gly410Arg | ENST00000403799.8:c.1228G>C | PM2 PP3 PM5 PP4 | Likely Pathogenic |
| 7:44146531:G:C | GCK | p.His317Gln | ENST00000403799.8:c.951C>G | PM2 PP3 PP1_Moderate PS4_Moderate PM5_Supporting PP4 PP2 | Likely Pathogenic |
| 7:44145176:G:A | GCK | p.Ser453Leu | ENST00000403799.8:c.1358C>T | PM2 PP3 PP1 PS4_Supporting PM5_Supporting PP4 PP2 PS3_Supporting | Likely Pathogenic |
| 7:44145212:G:A | GCK | p.Ser441Leu | ENST00000403799.8:c.1322C>T | PM2 PP3 PP2 PP4 PM5_Supporting | Likely Pathogenic |
| 7:44153372:C:A | GCK | p.Arg46Met | ENST00000403799.8:c.137G>T | PM2 PP3 PP2 PP4 PS4_Supporting | Pathogenic |
| 7:44149859:CCTGCCAAGAAGCA:C | GCK | p.? | ENST00000403799.8:c.580-13_580-1del | PVS1 PM2 PP1_Strong PP4 | Pathogenic |
| 7:44150978:A:G | GCK | p.Val154Ala | ENST00000403799.8:c.461T>C | PM2 PP3 PP1_Supporting PS4_Moderate PP4 PP2 | Likely Pathogenic |
| 7:44149992:G:A | GCK | p.Arg186* | ENST00000403799.8:c.556C>T | PVS1 PM2 PP1_Strong PS4 PP4 | Pathogenic |

| | | | | | |
|---|---|---|---|---|---|
| 7:44153299:A:G | GCK | p.? | ENST00000403799.8:c.208+2T>C | PVS1 PM2 PP3 | Pathogenic |
| 7:44146604:A:C | GCK | p.Ile293Arg | ENST00000403799.8:c.878T>G | PM2 PP3 PP1_Strong PS4_Moderate PM5 PP4 PP2 | Likely Pathogenic |
| 7:44152318:G:A | GCK | p.Gln106* | ENST00000403799.8:c.316C>T | PVS1 PM2 PP1 PS4_Moderate PP4 | Pathogenic |
| 7:44146619:C:T | GCK | p.? | ENST00000403799.8:c.864-1G>A | PVS1 PM2 | Pathogenic |
| 7:44147765:G:A | GCK | p.Arg250Cys | ENST00000403799.8:c.748C>T | PS4_Supplemntary PP1_Strong PP4_Moderate PM2 PP2 PP3 | Pathogenic |
| 7:44147741:C:T | GCK | p.Gly258Ser | ENST00000403799.8:c.772G>A | PM2 PP3 PS4_Moderate PM5_Supporting | Likely Pathogenic |
| 7:44145608:A:G | GCK | p.Met381Thr | ENST00000403799.8:c.1142T>C | PM2 PP3 PS4 PM5 PP1 | Pathogenic |
| 7:44149986:C:T | GCK | p.Ala188Thr | ENST00000403799.8:c.562G>A | PM2 PP1_Strong PS4 PM5_Supporting PP4 PP2 PS3 | Pathogenic |
| 7:44150954:A:G | GCK | p.? | ENST00000403799.8:c.483+2T>C | PVS1 PM2 PP3 PP4 PS4_Supporting | Pathogenic |
| 7:44145188:G:A | GCK | p.Ala449Val | ENST00000403799.8:c.1346C>T | PM2_Supporting PP2 PP3 PM5 PP4_Supporting | Likely Pathogenic |
| 7:44149779:G:T | GCK | p.Cys220* | ENST00000403799.8:c.660C>A | PVS1 PM2 PS4_Moderate PP1_Strong | Pathogenic |
| 7:44146463:C:A | GCK | p.Ser340Ile | ENST00000403799.8:c.1019G>T | PM2 PP3 PP1_Strong PS4_Moderate PM5 PP4 PP2 | Pathogenic |
| 7:44145576:G:A | GCK | p.Arg392Cys | ENST00000403799.8:c.1174C>T | PP2 PP3 PP4 PM2 PS4 PP1_Strong | Pathogenic |
| 7:44152268:CA:C | GCK | p.? | ENST00000403799.8:c.363+2del | PVS1 PM2 | Pathogenic |
| 7:44150008:A:C | GCK | p.Asn180Lys | ENST00000403799.8:c.540T>G | PM2 PP3 PS4 PP1_Strong | Pathogenic |
| 7:44146461:A:G | GCK | p.? | ENST00000403799.8:c.1019+2T>C | PVS1 PM2 PS3 PM3 PP1_Strong | Pathogenic |
| 7:44147720:C:T | GCK | p.Glu265Lys | ENST00000403799.8:c.793G>A | PM2 PP3 PS4_Moderate PS3 | Pathogenic |
| 7:44147690:G:C | GCK | p.Arg275Gly | ENST00000403799.8:c.823C>G | PM2 PP3 PM5 PP2 PP4_Supporting | Likely Pathogenic |
| 7:44145228:T:A | GCK | p.Ile436Phe | ENST00000403799.8:c.1306A>T | PM2 PP3 PS4 | Likely Pathogenic |
| 7:44150970:C:T | GCK | p.Glu157Lys | ENST00000403799.8:c.469G>A | PM2 PS4 | Likely Pathogenic |
| 7:44146619:C:G | GCK | p.? | ENST00000403799.8:c.864-1G>C | PVS1 PM2 PP3 | Pathogenic |
| 7:44153351:G:A | GCK | p.Ala53Val | ENST00000403799.8:c.158C>T | PM2 PP3 PP1_Strong PS4_Moderate PM5_Supporting PP4 PP2 | Pathogenic |
| 7:44151048:A:G | GCK | p.Ser131Pro | ENST00000403799.8:c.391T>C | PM2 PP1_Strong PP4 PP2 PS3_Supporting | Likely Pathogenic |
| 7:44147732:C:T | GCK | p.Gly261Arg | ENST00000403799.8:c.781G>A | PM2 PP2 PP3 PP4_Moderate PS3_Moderate PM3 PP1_Strong PS4 PS2 | Pathogenic |
| 7:44149968:C:T | GCK | p.? | ENST00000403799.8:c.579+1G>A | PVS1 PM2 | Pathogenic |
| 7:44152275:TCAGCAGTG:T | GCK | p.Thr118Aspfs*8 | ENST00000403799.8:c.351_358del | PVS1 PM2 PP1 PS4_Moderate PP4 | Pathogenic |

| | | | | | |
|---|---|---|---|---|---|
| 12:120994313:G:GC | HNF1A | p.Pro289Alafs*28 | ENST00000257555.11:c.863_864insC | PVS1 PS4_Moderate | Pathogenic |
| 12:120993584:G:T | HNF1A | p.Lys197Asn | ENST00000257555.11:c.591G>T | PM2 PS4_Supporting PP4_Moderate PP1_Supporting | Likely Pathogenic |
| 12:120988909:GA:G | HNF1A | p.Asp135Valfs*20 | ENST00000257555.11:c.404del | PVS1 PM2 PS4 PP1_Strong | Pathogenic |
| 12:120988898:G:A | HNF1A | p.Arg131Gln | ENST00000257555.11:c.392G>A | PM2 PP3 PM5 PS4_Moderate PS2 | Pathogenic |
| 12:120993591:C:T | HNF1A | p.Arg200Trp | ENST00000257555.11:c.598C>T | PS4 PP1_Strong PM2 PM5 PP3 PP4 | Pathogenic |
| 12:120994262:G:A | HNF1A | p.Arg271Gln | ENST00000257555.11:c.812G>A | PS4_Moderate PP1_Strong PM1 PP3 PM2 | Pathogenic |
| 12:120993519:G:A | HNF1A | p.? | ENST00000257555.11:c.527-1G>A | PVS1 PM2 PP3 PP4 PS4_Moderate PP1_Strong | Pathogenic |
| 12:120994267:AAAG:A | HNF1A | p.Glu275del | ENST00000257555.11:c.824_826del | PM2 PM4_Supporting PS4 PP1_Strong | Pathogenic |
| 17:37731599:C:CA | HNF1B | p.Ser348Valfs*12 | ENST00000617811.5:c.1040dup | PVS1 PM2 | Pathogenic |
| 20:44424117:G:T | HNF4A | p.Arg309Leu | ENST00000316673.8:c.926G>T | PM1_Supporting PM2 PP3 PM5_Supporting PP4_Moderate PP1_Moderate PS4_Moderate | Likely Pathogenic |
| 20:44413696:G:A | HNF4A | p.Val108Ile | ENST00000316673.8:c.322G>A | PM1_Supporting PM2 PP3 PS4_Moderate PP1_Strong PP4 | Pathogenic |
| 20:44406208:G:A | HNF4A | p.Arg67Gln | ENST00000316673.8:c.200G>A | PS4_Moderate PM2 PP3 PP1_Strong PP4_Moderate PM1 PM5_Supporting | Pathogenic |
| 20:44424116:C:T | HNF4A | p.Arg309Cys | ENST00000316673.8:c.925C>T | PS4 PM1_Supporting PP1_Moderate PP4_Moderate PP3 | Pathogenic |
| 20:44419741:C:T | HNF4A | p.Arg231Trp | ENST00000316673.8:c.691C>T | PP3 PM2 PM5 PS4_Moderate PP4 | Likely Pathogenic |
| 20:44413709:G:A | HNF4A | p.Arg112Gln | ENST00000316673.8:c.335G>A | PS4 PP1_Strong PP4_Moderate PM2 PP3 PM1 PM5_Supporting | Pathogenic |
| 20:44424117:G:A | HNF4A | p.Arg309His | ENST00000316673.8:c.926G>A | PM1_Supporting PM2 PP3 PM5_Supporting PP4 PP1_Moderate | Likely Pathogenic |
| 2:181678244:T:TG | NEUROD1 | p.His206ProfsTer38 | ENST00000295108.4:c.616dup | PVS1 PM2 PP1 PS4 | Pathogenic |
| 13:27920208:C:CCGGCGCCGGAGTT | PDX1 | p.Ser29GlyfsTer200 | ENST00000381033.5:c.72_84dup | PVS1 PM2 | Pathogenic |
| 13:27920410:C:CG | PDX1 | p.Leu92AlafsTer133 | ENST00000381033.5:c.273dup | PVS1 PM2 | Pathogenic |
| 13:27920342:C:A | PDX1 | p.Tyr68Ter | ENST00000381033.5:c.204C>A | PVS1 PM2 | Pathogenic |
| 13:27920443:AG:A | PDX1 | p.Gly103GlufsTer20 | ENST00000381033.5:c.308del | PVS1 PM2 | Pathogenic |
| 6:116882403:C:T | RFX6 | p.Arg181Trp | ENST00000332958.3:c.541C>T | PM2 PP3 PS4_Moderate PP4 PM3_Supporting | Likely Pathogenic |
| 6:116916217:T:G | RFX6 | p.Leu292* | ENST00000332958.3:c.875T>G | PVS1 PM2 PP1_Moderate PS4_Supporting | Pathogenic |

| | | | | | |
|---|---|---|---|---|---|
| 6:116877348:C:T | RFX6 | p.Gln25* | ENST00000332958.3:c.73C>T | PVS1 PM2 PS4_Moderate | Pathogenic |
| 6:116919165:A:T | RFX6 | p.Lys351* | ENST00000332958.3:c.1051A>T | PVS1 PM2 | Pathogenic |
| 6:116880544:G:A | RFX6 | p.Trp127* | ENST00000332958.3:c.381G>A | PVS1 PM2 | Pathogenic |
| 6:116925596:G:T | RFX6 | p.Gly608* | ENST00000332958.3:c.1822G>T | PVS1 PM2 | Pathogenic |
| 6:116919243:C:T | RFX6 | p.Arg377* | ENST00000332958.3:c.1129C>T | PVS1 PM2 PS4_Supporting PM3 | Pathogenic |
| 6:116894011:CAA:C | RFX6 | p.Lys198Argfs*26 | ENST00000332958.3:c.593_594del | PVS1 PM2 | Pathogenic |
| 6:116882404:G:A | RFX6 | p.Arg181Gln | ENST00000332958.3:c.542G>A | PM2 PP3 PM5_Supporting PS3 | Likely Pathogenic |
| 6:116925497:CT:C | RFX6 | p.Leu575Argfs*15 | ENST00000332958.3:c.1724del | PVS1 PM2 | Pathogenic |

**Supplemental Table 3: Pathogenic variants from the US health system–based Geisinger Mycode cohort identified in this study.**

| Genomic Change | Gene | Protein Change | NucleotideChange | Criteria | Classification |
|---|---|---|---|---|---|
| 11:17413396:G:A | ABCC8 | p.Arg826Trp | ENST00000302539.9:c.2476C>T | PM2 PP3 PS4_Strong PP4_Moderate PM3 PP1_Strong | Pathogenic |
| 7:44145602:G:A | GCK | p.Ser383Leu | ENST00000403799.8:c.1148C>T | PM2 PP3 PS4 | Likely pathogenic |
| 7:44146531:G:C | GCK | p.His317Gln | ENST00000403799.8:c.951C>G | PM2 PP3 PP1_Moderate PS4_Strong PM5_Supporting PP4 PP2 | Pathogenic |
| 7:44146565:A:G | GCK | p.Leu306Pro | ENST00000403799.8:c.917T>C | PM2 PP3 PP1 PS4_Supporting PM5_Supporting PP4 PP2 | Likely pathogenic |
| 7:44147649:C:T | GCK | - | ENST00000403799.8:c.863+1G>A | PVS1 PM2 | Pathogenic |
| 7:44147720:C:T | GCK | p.Glu265Lys | ENST00000403799.8:c.793G>A | PM2 PS4_Moderate PP4 PP2 PS3 | Pathogenic |
| 7:44147765:G:A | GCK | p.Arg250Cys | ENST00000403799.8:c.748C>T | PS4 PP1_Strong PP4_Moderate PM2 PP2 PP3 | Pathogenic |
| 7:44149763:C:T | GCK | p.Val226Met | ENST00000403799.8:c.676G>A | PS4 PP1_Strong PS3_Moderate PM2 PP2 PP4_Moderate PP3 PM1 | Pathogenic |
| 7:44149976:C:T | GCK | p.Arg191Gln | ENST00000403799.8:c.572G>A | PP3 PP2 PM2 PS4 PP1_Strong PP4_Moderate PM5_Supporting | Pathogenic |
| 7:44149982:A:G | GCK | p.Ile189Thr | ENST00000403799.8:c.566T>C | PM2 PP3 PP2 PS4 PP4_Moderate PP1_Strong | Pathogenic |
| 7:44150004:C:T | GCK | p.Val182Met | ENST00000403799.8:c.544G>A | PM2 PP1_Strong PS4 PM5_Supporting PP4 PP2 PS3 | Pathogenic |
| 7:44152338:C:T | GCK | p.Trp99Ter | ENST00000403799.8:c.296G>A | PSV1 PM2 | Pathogenic |
| 7:44153382:G:A | GCK | p.Arg43Cys | ENST00000403799.8:c.127C>T | PS4 PP1_Strong PP2 PP3 PP4 PM2 PS3_Moderate PM5 | Pathogenic |
| 7:44153387:A:C | GCK | p.Met41Arg | ENST00000403799.8:c.122T>G | PM2 PP3 PP1_Moderate PS4_Strong PM5_Supporting PP4 PP2 | Likely pathogenic |
| 7:44153411:A:G | GCK | p.Val33Ala | ENST00000403799.8:c.98T>C | PM2 PP3 PS4_Moderate PM5_Supporting PP4 PP2 | Likely pathogenic |
| 12:120988853:C:T | HNF1A | p.Ala116Val | ENST00000257555.11:c.347C>T | PM2 PP3 PP1_Strong PS4 | Pathogenic |
| 12:120988897:C:T | HNF1A | p.Arg131Trp | ENST00000257555.11:c.391C>T | PP3 PM1 PM2 PM5 PS4 PP1_Strong | Pathogenic |
| 12:120993601:G:A | HNF1A | p.Arg203His | ENST00000257555.11:c.608G>A | PM2_Supporting PP3 PS4 PP1_Moderate | Pathogenic |
| 12:120993619:C:A | HNF1A | p.Ala209Glu | ENST00000257555.11:c.626C>A | PS4_moderate PP3 PP4 PM1_Supporting PM2 | Likely pathogenic |
| 20:44419783:C:T | HNF4A | p.Arg245Cys | ENST00000316673.9:c.733C>T | PM2 PP3 PM5_Supporting PM6 PP4_Supporting | Likely Pathogenic |

| | | | | | |
|---|---|---|---|---|---|
| 20:44428356:A:AT | HNF4A | p.Ala363CysfsTer35 | ENST00000316673.9:c.1086dup | PVS1 PM2 | Pathogenic |
| 13:27920184:GA:G | PDX1 | p.Asp16AlafsTer107 | ENST00000381033.5:c.47del | PVS1 PM2 | Pathogenic |
| 13:27920230:GC:G | PDX1 | p.Pro33LeufsTer90 | ENST00000381033.5:c.98del | PVS1 PM2 | Pathogenic |
| 13:27920254:G:GCCGCCAGCCC | PDX1 | p.Pro45AlafsTer183 | ENST00000381033.5:c.122_131dup | PVS1 PM2 | Pathogenic |
| 13:27920308:T:TG | PDX1 | p.Glu58GlyfsTer167 | ENST00000381033.5:c.172dup | PVS1 PM2 | Pathogenic |
| 13:27920342:C:A | PDX1 | p.Tyr68Ter | ENST00000381033.5:c.204C>A | PVS1 PM2 | Pathogenic |
| 6:116882403:C:T | RFX6 | p.Arg181Trp | ENST00000332958.3:c.541C>T | PM2 PP3 PS4_Moderate PP4 PM3_Supporting | Likely pathogenic |
| 6:116919243:C:T | RFX6 | p.Arg377Ter | ENST00000332958.3:c.1129C>T | PSV1 PM2 PS4_Supporting PM3 | Pathogenic |
| 6:116919252:G:GT | RFX6 | p.Ser381IlefsTer22 | ENST00000332958.3:c.1139dup | PVS1 PM2 | Pathogenic |
| 6:116922040:A:AGAT | RFX6 | - | ENST00000332958.3:c.1329_1331dup | PVS1 PM2 | Pathogenic |
| 6:116927295:TC:T | RFX6 | p.Pro719LeufsTer54 | ENST00000332958.3:c.2156del | PVS1 PM2 | Pathogenic |
